# Heme induces mRNA expression and activation of tissue factor by TLR4 dependent mechanisms

**DOI:** 10.1101/2020.10.09.20210336

**Authors:** B.W. Hounkpe, C.R.P. Moraes, M.N.N. do Santos, F. F. Costa, E.V. De Paula

## Abstract

**Introduction:** Hemolytic diseases such as Sickle Cell Disease (SCD) are characterized by a natural propensity for both arterial and venous thrombosis. Evidence showing that heme can induce tissue factor (TF) expression in endothelial cells and TF-dependent coagulation activation in animal models of SCD suggest that heme can contribute to hypercoagulability in this condition. We recently demonstrated that heme can induce coagulation activation in whole blood of healthy volunteers in a TF-dependent fashion.

**Methods:** Herein, we aimed to evaluate whether this heme-induced coagulation activity was dependent on the expression and/or activation of hematopoietic TF in human mononuclear cells. *TF* mRNA expression was evaluated by qPCR and TF procoagulant activity was evaluated using a 2-stage assay based on the generation of FXa.

**Results:** Heme was capable of inducing *TF* expression and activation in a TLR4-dependent pathway. This activity was further amplified after TNF-α-priming.

**Conclusion:** Our results provide additional evidences on the mechanisms by which heme is involved in the pathogenesis of hypercoagulability in hemolytic diseases.

## Introduction

Clinical and experimental data clearly demonstrate that hemolytic anemias such as Sickle Cell Disease (SCD), β thalassemia, paroxysmal nocturnal hemoglobinuria and autoimmune hemolytic anemia are associated with hypercoagulability ^1–3^. Heme is an essential molecule present in almost all forms of life. However, evidence gathered in the last two decades show that free heme, which can be released from hemoglobin during intravascular hemolysis, can be toxic to cells by both direct (via Fenton reaction and reactive oxygen species generation) and indirect (innate immunity-mediated) mechanisms ^4^. The effects of heme on several compartments of innate immunity have been described over the last two decades, leading to the concept that free heme can behave as an erythrocyte danger associated molecular pattern (DAMP), through the activation of TLR4 receptors ^5,6^. Since hemostasis is currently regarded as part of innate immunity ^7,8^, heme has also been studied as a potential mediator of hypercoagulability.

In this regard, heme has been shown to induce the expression of tissue factor (TF) in cultured endothelial cells ^9^, and was subsequently characterized as a mediator of coagulation activation *in vivo* in mice models of SCD, in a study that also demonstrated that heme was capable to activate TF in human monocytes ^10^. Moreover, heme has also been shown to induce the formation of neutrophil extracellular traps, which facilitate coagulation activation during thromboinflammation ^11^. Recently, we demonstrated that incubation of heme in whole blood from healthy volunteers triggers coagulation activation in a TF-dependent fashion ^12^. Together, these results support the concept that free heme could be involved in the initiation and/or maintenance of hypercoagulabity in SCD and other hemolytic conditions.

Yet, the mechanisms by which heme activates hemostasis are not entirely clear, and limited evidence is available supporting its effect on TF mRNA expression and activation. Here we characterized in more detail the effect of heme on TF expression and activation in human peripheral blood mononuclear cells (PBMCs), and explored the effects of TNF-α and TLR-4 in this process.

## Methods

### Obtention of PBMC from blood donors

This study was performed in accordance with the Declaration of Helsinki and approved by the Ethics Committee of University of Campinas (CAAE: 46853115.4.0000.5404). Peripheral blood mononuclear cells (PBMCs) were obtained from buffy coats as previously described ^13,14^, using de-identified blood bags provided after whole blood fractionation from the blood center of University of Campinas (Hemocentro). Bags containing residual buffy coats were conserved at 4° C overnight and quickly pre-processed the next day. Peripheral blood mononuclear cells (PBMCs) were isolated with 1.077 density of ficoll (Ficoll-paque Plus, GE Healthcare) after dilution of the obtained buffy coat with PBS 1X (1:3 dilution vol:vol). After separation of PBMCs, cells were seeded in culture plates at a quantity equivalent to 5*10^6^ and 2*10^6^ monocytes (based on automated cell counts in a hematology analyzer) for quantitative PCR (qPCR) and functional coagulation assays, respectively. Cells were then incubated at 37°C for 4h with heme or vehicle at different concentrations. When necessary, cells were primed with 2ng/mL of TNF-α at 37°C for 1h or pre-treated with 5ng/ml of TLR4 specific inhibitor Tak-242. After incubation, supernatants were removed to harvest only adherent cells in order to enrich the monocyte content of the sample ^15–18^. Cells were washed with PBS and re-suspended in HBSA buffer to measure TF pro-coagulant activity (TF PCA) with a two-stage assay based on FXa generation, or were lysed with Trizol for *TF* mRNA expression by qPCR. Pre-processed samples were stored at −80°C until total RNA extraction or TF PCA analysis.

### Whole blood collection and plasma separation

Whole blood samples were collected from healthy volunteers by venipuncture in vacuum tubes containing 3.2% sodium citrate (BD Vacutainer Coagulation Tube, 2.7 ml, 0.109 M buffered sodium citrate). These samples were also treated with heme or vehicle. Samples were then incubated at 37°C for 4h. Plasma samples were immediately processed to perform one-stage coagulation assay.

### Heme dilution

Heme (Ref H651-9; Frontier Scientific, USA) was diluted with 100μl of NaOH 0.1M then adjusted with 900 μl of ultra-filtered water to initial working concentration of 5 mM. This solution was filtrated though a 0.22μm filter and freshly used according the scenario.

### Gene expression quantification by qPCR

Total RNA was extracted using Trizol using classical methods. For each sample, 1μg of RNA was transcribed using High-capacity cDNA Reverse Transcription Kit (Applied Biosystems™). *TF* gene expression was evaluated by qPCR and normalized using the geometric mean of two reference transcripts EEF2 (ENST00000309311) and MYH9 (ENST00000216181) provided by HT Atlas database ^19^. These transcripts were shown to be among the most stably expressed in PBMCs and have been validated in our experiment.

### Two-stage TF PCA assay

TF PCA of adherent fractions of PBMCs were evaluated by a two-stage chromogenic assay based on the kinetics of FXa generation, as previously described. Briefly, 30μl of treated cells suspended in HBSA was incubated with 20μl of HBSA containing 150nM FX, 1nM FVIIa and 3mM CaCl_2_ for 2 hours at 37°C ^20^. In these experiments, an IgG control antibody and an anti-TF antibody treatment (15 min of incubation at room temperature with 400μg/ml final concentration) were used to measure the fraction of Xa generation that is dependent on TF. After incubation, FXa generation was stopped by 25 μL of EDTA buffer (25 mM) for 10 min, and then 25 μL of 0.3mM FXa substrate Pefachrome sFXa (Pefachrome®) was added for absorbance reading at 405 nm every 30 seconds, for 30 minutes. Absorbance was referred to an Innovin (2.5nM-0.02nM) standard curve. Final results were expressed as the equivalent of TF (in pg/mL) to the procoagulant activity from IgG treated samples subtracted from procoagulant activity in anti-TF treated samples.

### One-stage clotting test

Heme-induced procoagulant activity of plasma was measured using a one-stage clotting test performed using a semi-automated coagulometer (Max Start 4, Diagnostica Stago) after pre-incubation of heme at 37°C with normal human plasma, followed by re-calcification as previously described ^21^. The procoagulant activity was determined using standard curve generated with a recombinant human relipidated TF (Innovin, Dade Behring). Results were reported in units of TF activity. In 50 μl of test sample, one unit of TF activity was arbitrarily defined as the amount of TF which yields a 50 seconds clotting time with pooled normal plasma ^22^.

### Statistical analysis

Experiments were repeated at least 3 times, and results were expressed as the median of each variable. Statistical analysis was performed using GraphPadPrism 7.0 (GraphPadPrism Software Inc. San Diego, California, USA). Wilcoxon two-side or Friedman tests were used to compare quantitative parameters obtained from the same samples exposed to vehicle or heme, with or without TNF-α. Correction for multiple comparisons was performed using Dunn’s test. A p-value of < 0.05 was considered significant.

## Results and discussion

### Heme induces TF gene expression in PBMCs

Heme has been previously shown to induce procoagulant activity in whole blood ^12^, and to activate TF activity in monocytes ^10^. Here we explored whether this effect was dependent on hematopoietic *TF* gene expression. When compared to vehicle, heme significantly induced *TF* gene expression measured by qPCR in PBMC after 4h of incubation (Figure 1). The induction was evident with heme concentrations above 10μM, suggesting a dose dependent effect.

**Figure 1:**
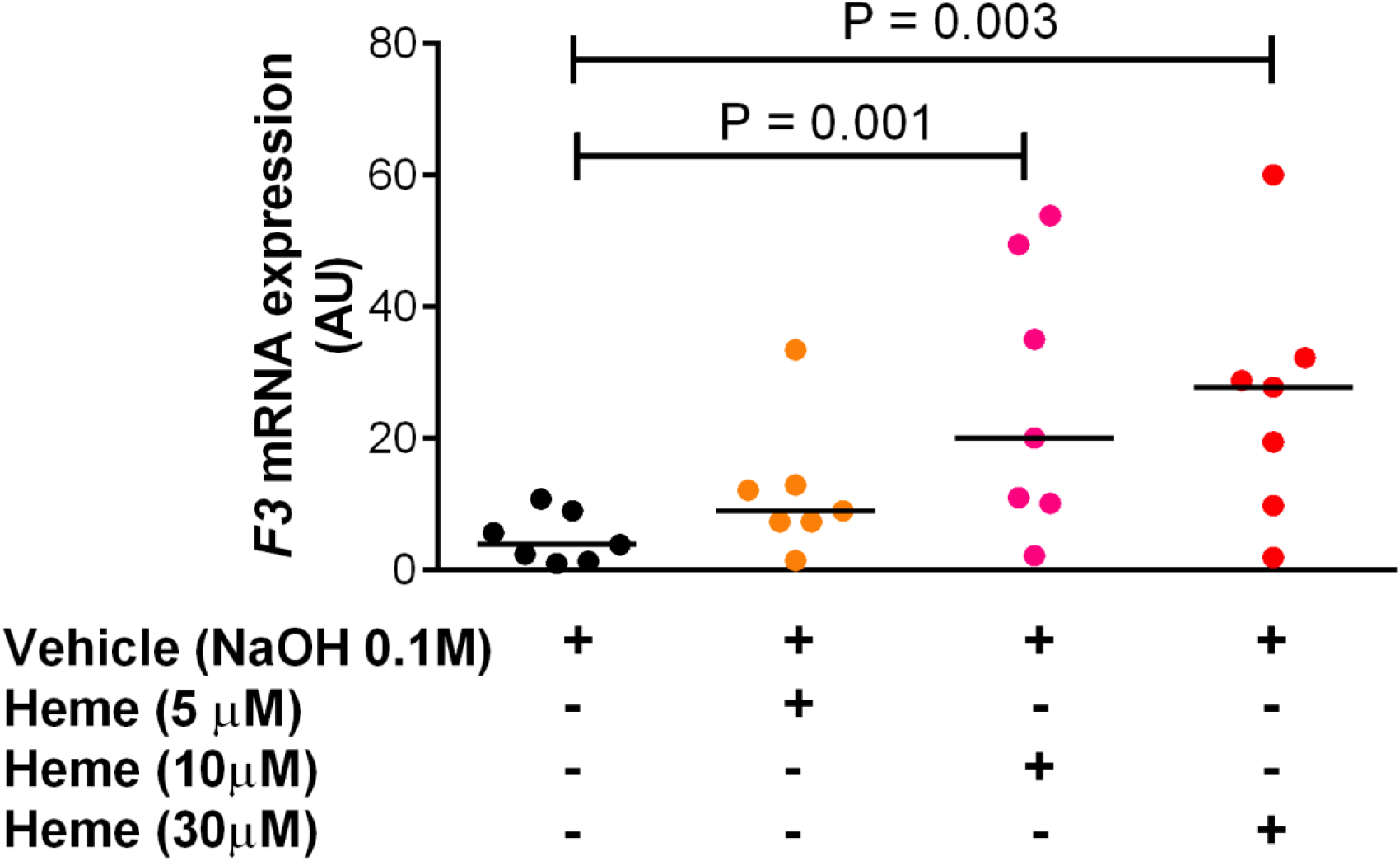
Evaluation of *F3* mRNA expression in mononuclear cells by qPCR. PBMCs were used to perform qPCR after 4h of incubation with vehicle and heme at different concentrations. Results were expressed in arbitrary units (AU) as median of normalized *F3* expression using two stably expressed reference transcripts *EEF2* (ENST00000309311) and *MYH9* (ENST00000216181). Comparisons were made between vehicle and increasing concentrations of heme and showed that heme induces TF mRNA expression in a dose response fashion. Friedman test was performed and multiple comparisons were corrected using the Dunn’s test; a p-value of < 0.05 was considered significant; seven samples were analyzed.

### Heme triggers TF procoagulant activity

We next explored the effects of heme on TF activation. Heme 30μM induced a significant increase in TF procoagulant activity of PBMCs when compared to vehicle, and this effect was enhanced by priming cells with TNF-α prior to heme stimulation (Figure 2A). We also demonstrated that heme-induced TF activation seems to be dose dependent (Figure 2B). TNF-α alone at a concentration of 2ng/ml was not able to induce TF procoagulant activity.

**Figure 2:**
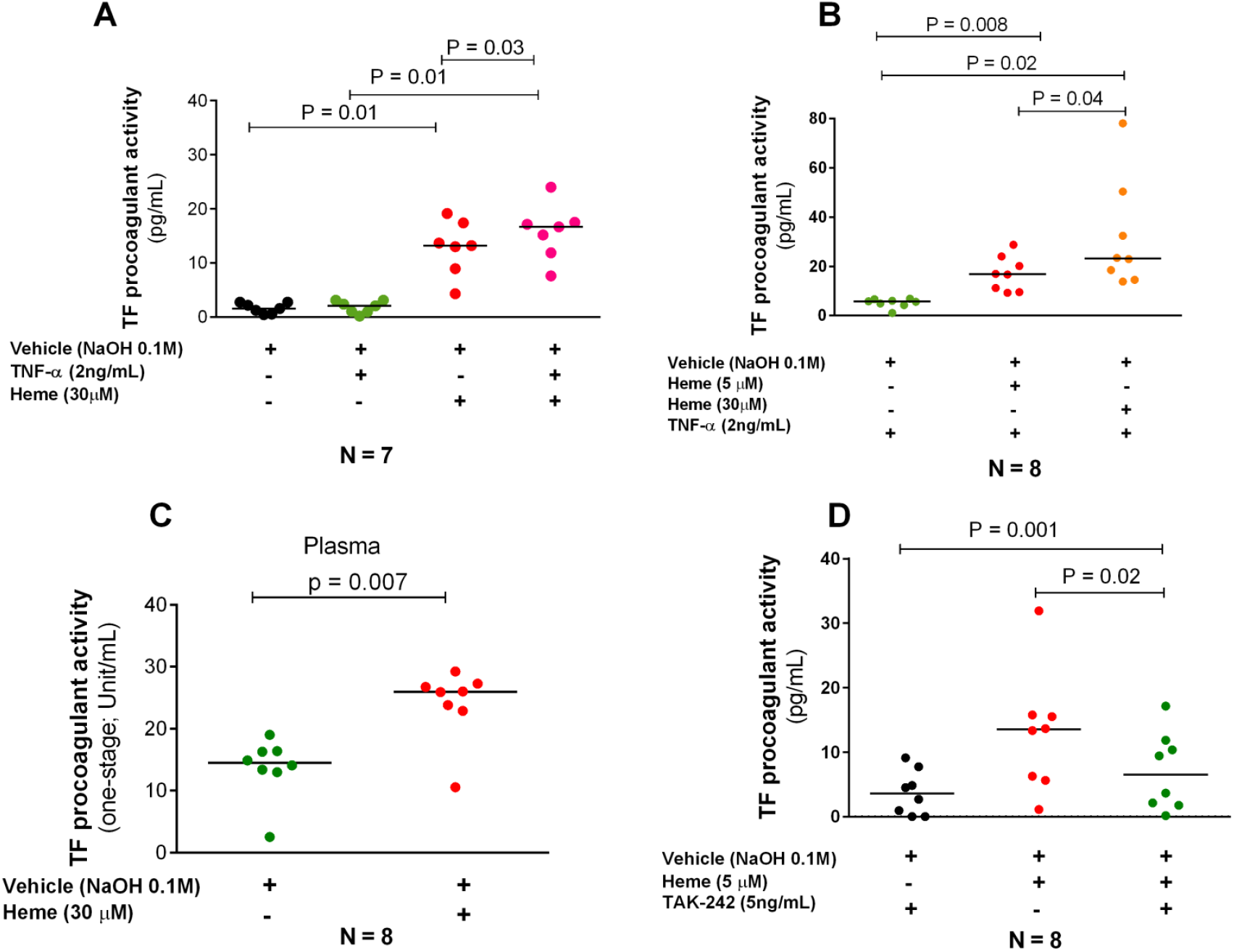
Heme-induced procoagulant activity is dependent of TLR4 signaling and potentiated by TNF-α. Comparisons were made for heme *vs* vehicle. When indicated cells were primed with TNF-α. Results were ‘presented as median of procoagulant activity in ng/ml. Heme-induced TF procoagulant activity is potentiated by TNF-α-priming (A) as shown by the comparison between heme (30 μM) and heme (30 μM) + TNF-α; this heme effect is also dose dependent (B); heme pro-coagulant effect was then confirmed in plasma using a coagulometric assays (C) illustrating the relevance of circulating TF and plasma components; Tak-242, an inhibitor of TLR4 reverted heme-induced TF PCA (D). Data are presented as median of TF procoagulant activity (A,B and D) or units of TF activity (C). Wilcoxon two-tail test was performed to compare the variables and sample size was indicated in each panel.

Since circulating TF and plasma components such as micro-vesicles carrying heme have been shown to be relevant to coagulation activation ^23,24^ we hypothesized that heme might also be capable of inducing procoagulant activity in plasma. Accordingly, using a one-stage coagulometric assay we were able to show that heme induces procoagulant activity in plasma separated from whole blood after 4 hours of incubation with heme (Figure 2C).

### TF activity induced by heme is dependent on TLR4

Heme is an erythrocyte DAMP and was shown to be a potent inducer of inflammation and to trigger of vaso-occlusive events in animal models of SCD via TLR4 receptors ^5,25,26^. Accordingly, we explored whether heme triggers TF procoagulant activity through TLR4 by using Tak-242, which was capable to revert heme-induced TF PCA (Figure 2D).

SCD is characterized by increased risk of venous thromboembolism ^27^. Heme has been implicated as a mediator of hypercoagulability in SCD by indirect evidence of its effects on innate immunity, and by a study in an animal model of SCD, which showed that heme effects are mediated by TF activation in mice ^10^. In order to gain insights into the mechanisms of hypercoagulability in SCD and other hemolytic anemias, we explored additional details of heme-induced coagulation activation in human mononuclear cells. In this context, the most important results of our study were the demonstration that heme is capable of inducing both mRNA expression and activation of hematopoietic TF in a dose-dependent and TLR-mediated fashion.

Using a global hemostasis assay, we recently demonstrated that heme was capable of coagulation activation in whole blood and that this effect was more related to the kinetic of clot formation than its intensity ^12^. Although it has been demonstrated that heme can trigger TF expression in endothelial cells, studies using endotoxemia models have shown that hematopoietic TF is more relevant for coagulation activation ^28^, and in fact heme has been previously shown to active TF in monocytes ^10^. Here we provide additional details about the effects of heme on the expression and activation of hematopoietic TF, confirming it using two different coagulometric assays, and that these effects are dose-dependent.

Pathogenesis of hypercoagulability in SCD is multifactorial and involves crosstalk with other inflammatory pathways ^3,29^. Accordingly, we also demonstrated that heme-induced TF PCA was potentiated by TNF-α. Of note, synergy of heme with other pro-inflammatory mediators have been previously described in experiments involving LPS stimulation ^30^ and in an *in vitro* model of NETosis in neutrophils pretreated with TNF-α ^11^. However, as far as we are aware, there are no reports of a priming effect of TNF-α in TF activation. Moreover, similar to what is described with other compartment of innate immunity ^11,26^, we also demonstrated that heme-induced TF activation is TLR-dependent.

In conclusion, we demonstrate that heme, an ubiquitous molecule that is released in several clinical conditions associated with hemolysis can trigger both TF mRNA expression and TF activation in human hematopoietic cells, and that this effect was dependent on TLR4 receptors. These results reinforce the concept that free heme may play a relevant role in the pathogenesis of hypercoagulability in SCD, as well as in other hemolytic conditions whose physiopathology involve hypercoagulability.

## Data Availability

All data generated during this study are included in this article.

## Author contributions statement

BWH and CRPM performed experiments; BWH analyzed, interpreted data and drafted the manuscript; MNNS interpreted data; FFC and EVP contributed with reagents and laboratory infrastructure; EVP designed the study, reviewed and analyzed data and drafted the manuscript. All authors revised and approved all submitted versions of the manuscript.

